# Macrolide and non-macrolide resistance after 36 months of mass azithromycin distribution in Burkina Faso: A cluster randomized trial

**DOI:** 10.1101/2025.08.14.25333693

**Authors:** Catherine E. Oldenburg, Boubacar Coulibaly, Ali Sié, Mamadou Ouattara, Mamadou Bountogo, Guillaume Compaoré, Dramane Kiemde, Adama Compaoré, Guillaume Zonou, Armin Hinterwirth, Lina Zhong, Cindi Chen, YuHeng Liu, Danny Yu, Thomas Abraham, Elodie Lebas, Huiyu Hu, Milan Hilde-Jones, Benjamin F. Arnold, Thuy Doan, Thomas M. Lietman

## Abstract

**Background:** Biannual mass azithromycin distribution to children aged 1-59 months reduces all-cause child mortality but is known to select for antimicrobial resistance (AMR). The World Health Organization (WHO) recommends ongoing surveillance of AMR in the context of azithromycin distribution. Here, we evaluated the impact of twice-yearly distribution of azithromycin compared to placebo on AMR in the gut of children participating in a trial in Burkina Faso.

**Methods:** The Community Health with Azithromycin Trial (CHAT) was a 1:1 cluster randomized placebo-controlled trial in Nouna District, Burkina Faso from 2019-2023. Communities were randomized in a 1:1 fashion to twice yearly azithromycin (20 mg/kg) or matching placebo. At 36 months, rectal swabs were collected from a random sample of 15 children per community in 48 communities participating in the trial and assessed for AMR genetic resistance determinants using DNA-seq. We assessed the fold-change in macrolide and non-macrolide resistance determinants between treatment groups after 36 months.

**Results:** 483 samples from 41 communities were analyzed at 36 months. Macrolide resistance determinants were not statistically significantly higher in the azithromycin group compared to the placebo group (fold-change 1.21, 95% confidence interval, CI, 0.96 to 1.52, *P*=0.62). There was no evidence of a difference in non-macrolide resistance genes, for example, beta-lactam resistance was similar between treatment groups (fold-change 1.05, 95% CI 0.79 to 1.40, *P*=0.81).

**Conclusions:** In this setting in Burkina Faso, twice-yearly azithromycin distributions to children aged 1-59 months did not lead to statistically significant differences in macrolide or non-macrolide genetic resistance determinants at 36 months.

**Trial Registration:** ClinicalTrials.gov NCT03676764

**KEY POINTS:** Twice-yearly mass distribution of azithromycin to children aged 1-59 months did not lead to a statistically significant difference in genetic macrolide or nonmacrolide resistance determinants in the gut after 36 months of distributions.

## INTRODUCTION

Twice yearly mass distribution of azithromycin has been shown to reduce all-cause mortality by 14-18% in children aged 1-59 months in Burkina Faso and Niger.^1–3^ Mass distribution of azithromycin to all members of a community for trachoma control has been shown to lead to community-level increases in the prevalence of macrolide resistance in *Streptococcus pneumoniae* and other potentially pathogenic organisms, although the prevalence of resistance generally returns to baseline levels once antibiotic selection pressure is removed.^4–8^ In the *Macrolides Oraux pour Réduire les Décès avec un Oeil sur la Résistance* (MORDOR)-Niger study, a statistically significant increase in genetic macrolide resistance determinants was documented after 36 and 48 months of azithromycin distributions compared to placebo, with approximately a 7.5-fold increase in macrolide resistance in azithromycin communities compared to placebo.^9^ In addition to macrolide resistance, this study found some evidence of an increase in beta-lactam resistance in communities receiving azithromycin compared to placebo, suggesting the potential for co-selection of resistance determinants.^9^

The MORDOR-Niger study was done in an area that was not receiving seasonal malaria chemoprevention (SMC), which typically consists of 3-5 monthly rounds of mass drug administration with sulfadoxine-pyrimethamine (SP) and amodiaquine (AQ) to children aged 3-59 months. A household randomized study of SMC distributed with or without azithromycin demonstrated little evidence of selection for resistance in pneumococcus in households receiving azithromycin compared to placebo in Burkina Faso.^10^ Sulfadoxine has bacteriostatic properties, and it is possible that distribution of SP changes antimicrobial resistance selection dynamics for azithromycin. Here, we evaluated selection for macrolide and nonmacrolide genetic resistance determinants in the Community Health with Azithromycin Treatment (CHAT) trial in communities that had received 36 months of twice-yearly azithromycin or placebo to children aged 1-59 months in Nouna District, Burkina Faso and in which an 18% reduction in mortality was observed in communities receiving azithromycin compared to placebo.^1,11^ We hypothesized that twice yearly mass distribution of azithromycin would lead to selection for macrolide but not non-macrolide genetic resistance determinants in a setting where SMC was being distributed from July through October.

## METHODS

### Trial design

Complete methods for the CHAT trial have been previously reported.^1,11^ CHAT was a 1:1 community randomized, placebo-controlled trial designed to evaluate whether a twice-yearly distribution of azithromycin to children aged 1-59 months reduced mortality compared to matching placebo in Nouna District, Burkina Faso. The primary outcome has previously been reported.^1^ A subset of 48 randomization units in the Nouna Health and Demographic Surveillance Site (HDSS)^12^ were randomly selected at baseline for inclusion in morbidity monitoring as part of the trial. The HDSS covers approximately 1/3 of Nouna District. All communities in Nouna District were eligible for the trial, but eligibility for the morbidity monitoring was restricted to those in the HDSS due to the proximity to the laboratory and resources for sample collection. In communities selected for morbidity monitoring, rectal swabs were collected from a random sample of 15 children per randomization unit at baseline and 36 months. Baseline sample collection was conducted from August through November 2019.

Endline sample collection occurred at 36 months (December 2022 through February 2023), approximately 6 months following the final azithromycin distribution. At the 36-month measurement, some areas of Nouna District were inaccessible due to insecurity, and thus we were unable to include all the original clusters. We randomly selected 16 replacement clusters that were participating in the trial but were not included in baseline morbidity monitoring to increase statistical power. The trial was reviewed and approved by the Institutional Review Board at the University of California, San Francisco and the Comité e’Ethique pour la Recherche en Santé in Ouagadougou, Burkina Faso. Written informed consent was obtained from the caregiver of each child enrolled in the trial.

### Randomization, interventions, and masking

Communities were randomized in a 1:1 fashion stratified by whether they were in the HDSS. Communities were randomized to receive a single, oral dose of 20 mg/kg azithromycin or matching placebo delivered via door-to-door census at approximately 6-monthly intervals (5 distributions total prior to final sample collection).

Azithromycin and placebo were identical in composition but excluded the active ingredient. Participants, caregivers, study staff, and investigators were masked to treatment assignment. Laboratory personnel were masked to treatment assignment and to randomization unit.

### Sample collection

Rectal swabs were collected at baseline and 36 months and placed in the Zymo stool collection kit (Zymo Research, Irvine, CA) for nucleic acid preservation. Samples were stored at −80°C at the laboratory in Nouna until they were shipped to the United States for processing.

### Laboratory methods

Rectal samples were processed for DNA-seq as previously described.^13^ Briefly, rectal samples were pooled at the community level and extracted using the Maxwell RSC Enviro TNA Kit (Promega, Madison, WI, United States) per manufacturer’s instructions and prepared for DNA sequencing libraries using the New England Biolabs’ (NEB, Ipswich, Massachusetts, United States of America) NEBNext Ultra II DNA Library Prep Kit and then amplified with 10 PCR cycles and sequenced on the NovaSeq X (Illumina) using 150-nucleotide (nt) paired-end sequencing. Sequencing reads were host-read removed and quality filtered.

Remaining reads were aligned to the MEGARes reference antimicrobial database (version 3.0.0) using BWA with default settings.^14,15^ Only antimicrobial-resistant genes (ARGs) were included if they met 80% nucleotide identity between the query and the reference AMR sequence. Any hits that failed SNP confirmation (for genes that required it) were excluded. Each ARG then was grouped at the class level (such as macrolides, β-lactams, aminoglycosides, etc.) using Resistome Analyzer.

### Statistical analysis

For comparisons of macrolide resistance gene load at 36 months, we anticipated that 48 clusters (N=24 per arm) with 15 children per cluster would yield approximately 80% power to detect an absolute difference of 0.082 in mean normalized genetic load between azithromycin and placebo arms (approximately 1.16-fold difference), assuming a placebo resistance prevalence of 0.05 and an intracluster correlation coefficient of 0.1.

Differential abundance analysis of AMR gene classes between azithromycin and placebo arms at 36 months post-treatment was performed using DESeq2 (version 1.46.0), incorporating normalization by the number of non-host (non-human) sequencing reads per sample.^16^ To ensure statistical stability, AMR genes were filtered to include only those with at least 8 non-zero observations across all samples, and with counts detected in both treatment arms. A negative binomial generalized linear model was fitted to the normalized counts, with treatment arm as the predictor. Fold changes in AMR gene abundance were estimated using DESeq2’s empirical Bayes shrinkage procedure, and Wald tests were used to assess statistical significance. Adjusted p-values were calculated across non-macrolide antibiotic classes using the Benjamini–Hochberg method to control the false discovery rate (FDR) at 5%. Individual antimicrobial resistance gene (ARG) - level analysis for macrolide resistance genes followed the same DESeq2 workflow and p-value adjustment. In addition, we calculated alpha diversity metrics, including Shannon index and inverse Simpson index, using bacteria genus-level profiles and compared these between treatment arms using t-tests. Diversity measures were calculated in the R package “vegan.” All analyses were conducted in R (version 4.5.0).

## RESULTS

At baseline, a total of 615 children in 41 communities were sampled; 1 community in the azithromycin group and 6 in the placebo group that had been randomly selected for inclusion in morbidity monitoring were inaccessible due to insecurity and were not included in the baseline assessment (**Figure 1**). At 36 months, 7 communities in each group were inaccessible due to insecurity, and 6 replacement communities were included in the azithromycin group and 10 from the placebo group were included. Samples from 2 communities at 36 months were lost during sample transport. At 36 months, a total of 483 samples from 41 communities were analyzed. At baseline, approximately half of the children in each treatment group were female, and the age distribution was similar between groups (**Table 1**). Genetic macrolide resistance determinants were slightly higher in the azithromycin group compared to the placebo group at baseline (**Table 1**).

**Figure 1.**
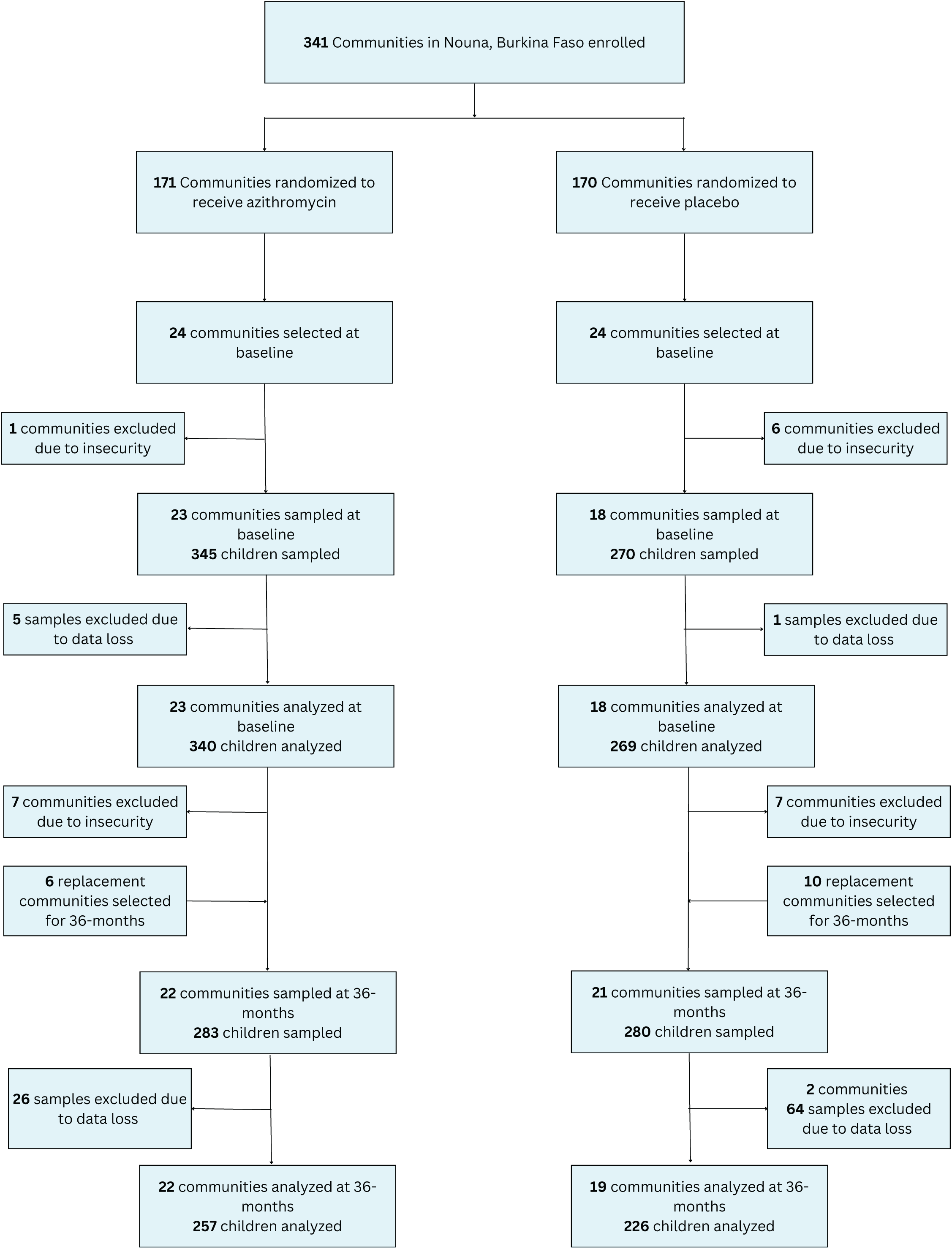
CONSORT flow diagram of study participation.

**Table 1.**
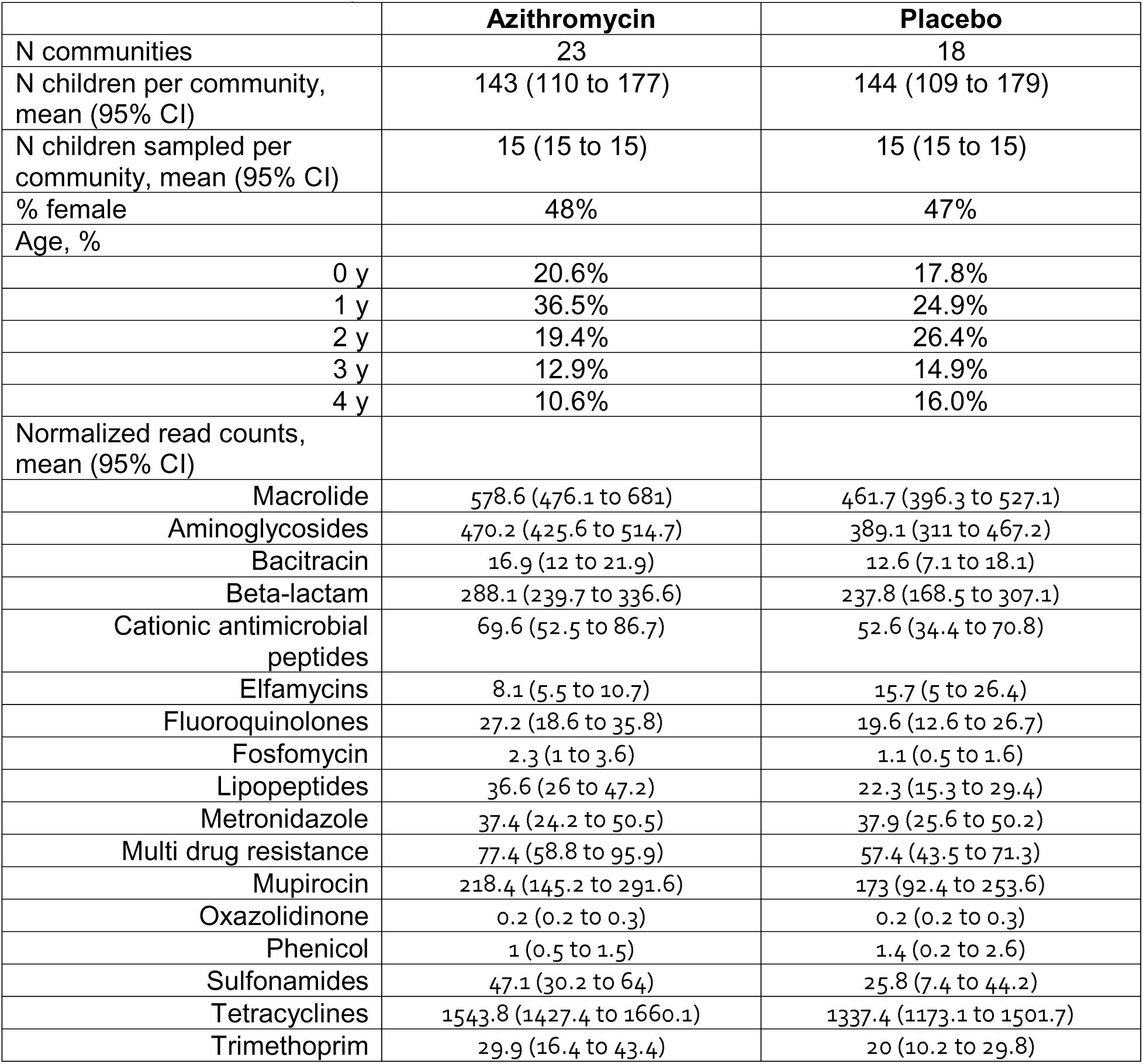
Baseline community and child characteristics.

We found no evidence of a difference at the class level for antibiotic resistance determinants at 36 months between azithromycin and placebo communities. There was a non-statistically significant 1.21-fold increase in macrolide resistance determinants in azithromycin-treated communities compared to placebo (95% confidence interval, CI, 0.96 to 1.52, *P*=0.62, **Figure 2**). There was no evidence of a difference in beta-lactam resistance between groups (fold change 1.05, 95% CI 0.79 to 1.40, *P*=0.81; **Figure 2A**) or to any other class of antibiotics (**Figure 2A**). At the gene level, there was an increase in *ermF* in communities receiving azithromycin (fold change 2.21, 95% CI 1.08 to 4.50; **Figure 2B**), but this difference was not statistically significant after accounting for multiple comparisons (*P*=0.32). There was no evidence of a difference in fold change in any other ARG.

**Figure 2.**
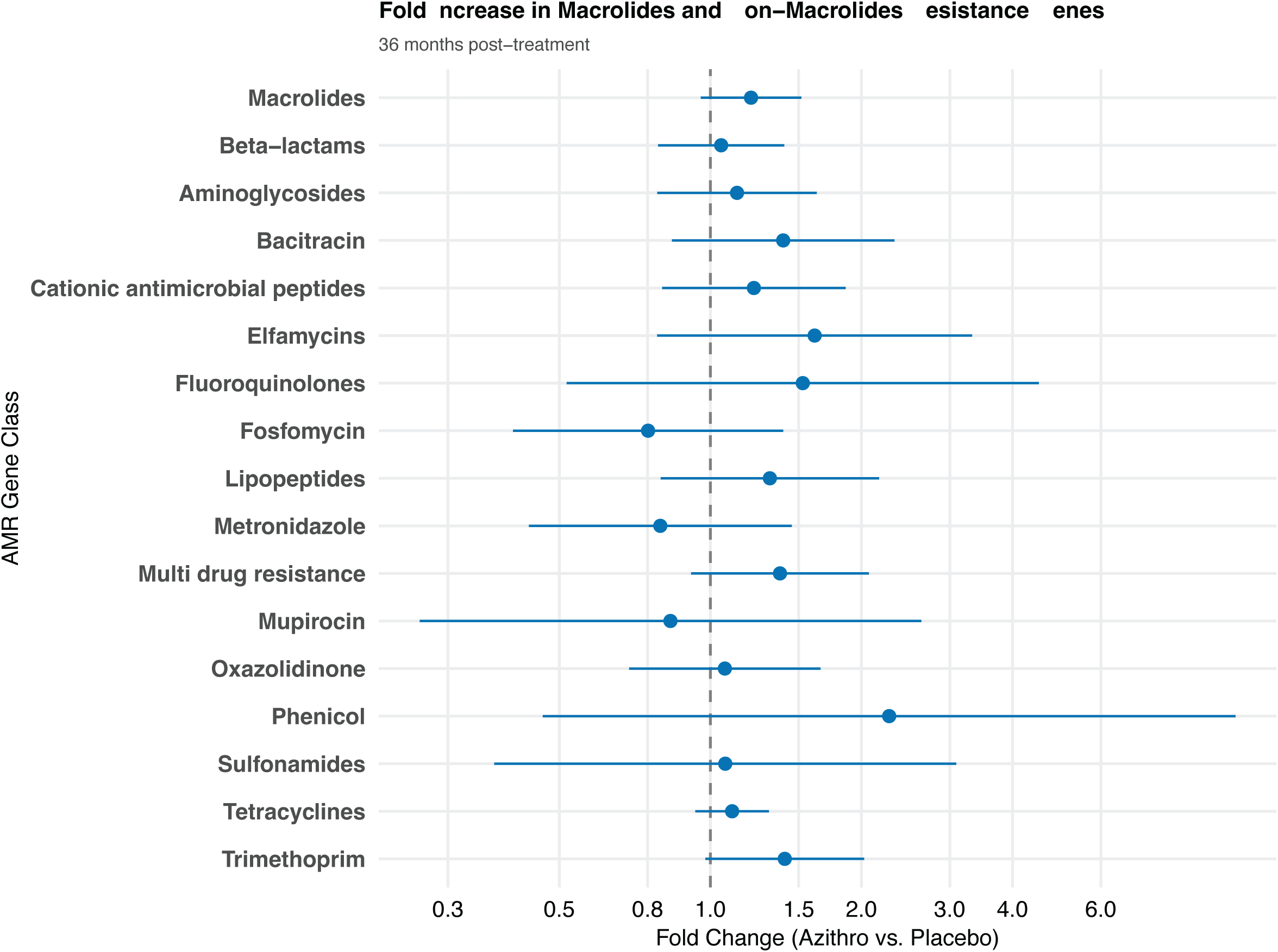
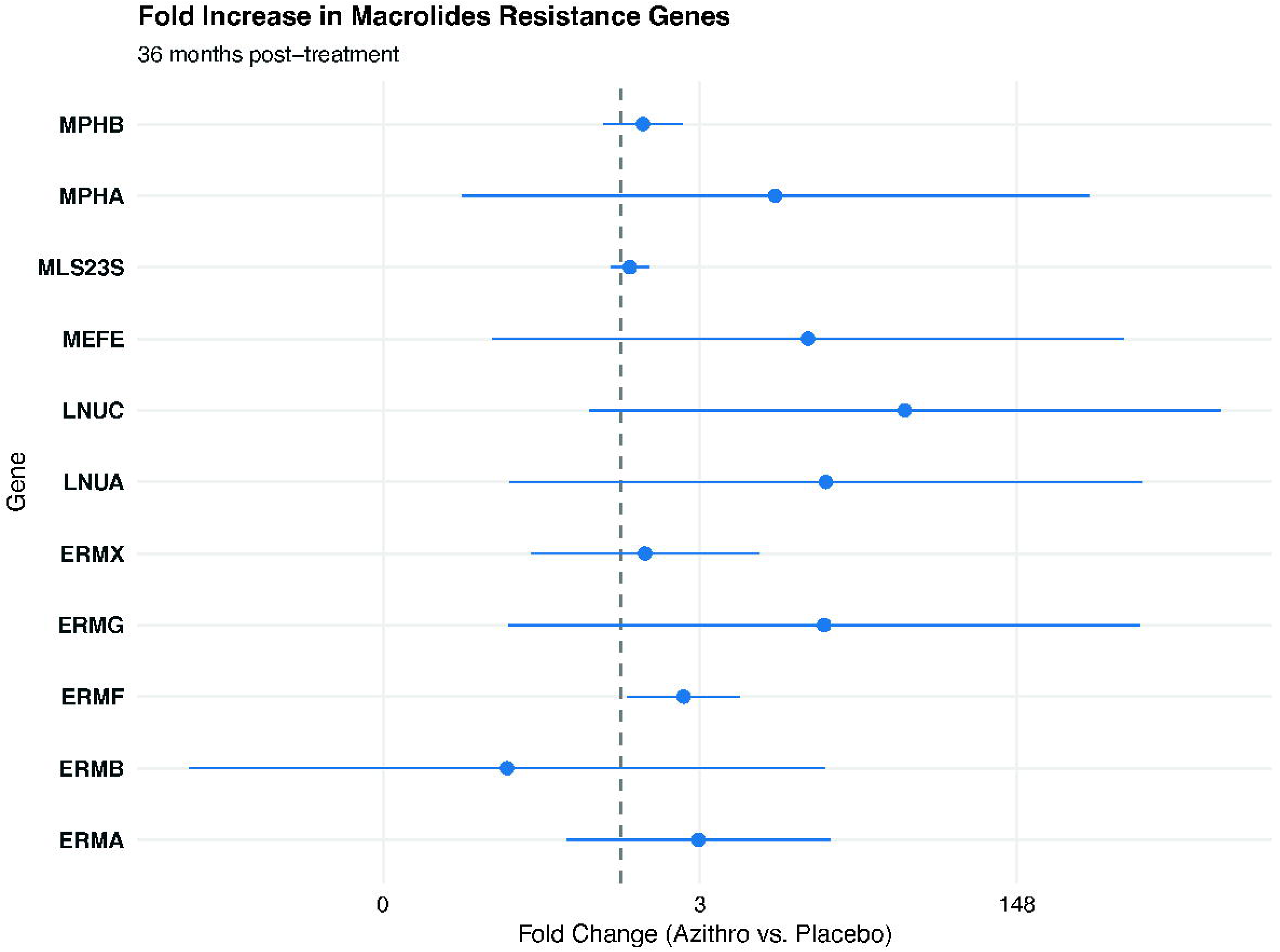
Fold increase in macrolide and non-macrolide resistance genes at the class level (A) and individual macrolide antibiotic resistance genes (B) at 36 months in communities receiving twice-yearly azithromycin distribution compared to placebo.

At the bacterial genus level, Shannon’s index and inverse Simpson’s index were similar between the two treatment groups, with no evidence of a difference in microbiome diversity between groups (Shannon’s: *P*=0.76, **Figure 3A**; Simpson’s: *P*=0.64, **Figure 3B**).

**Figure 3.**
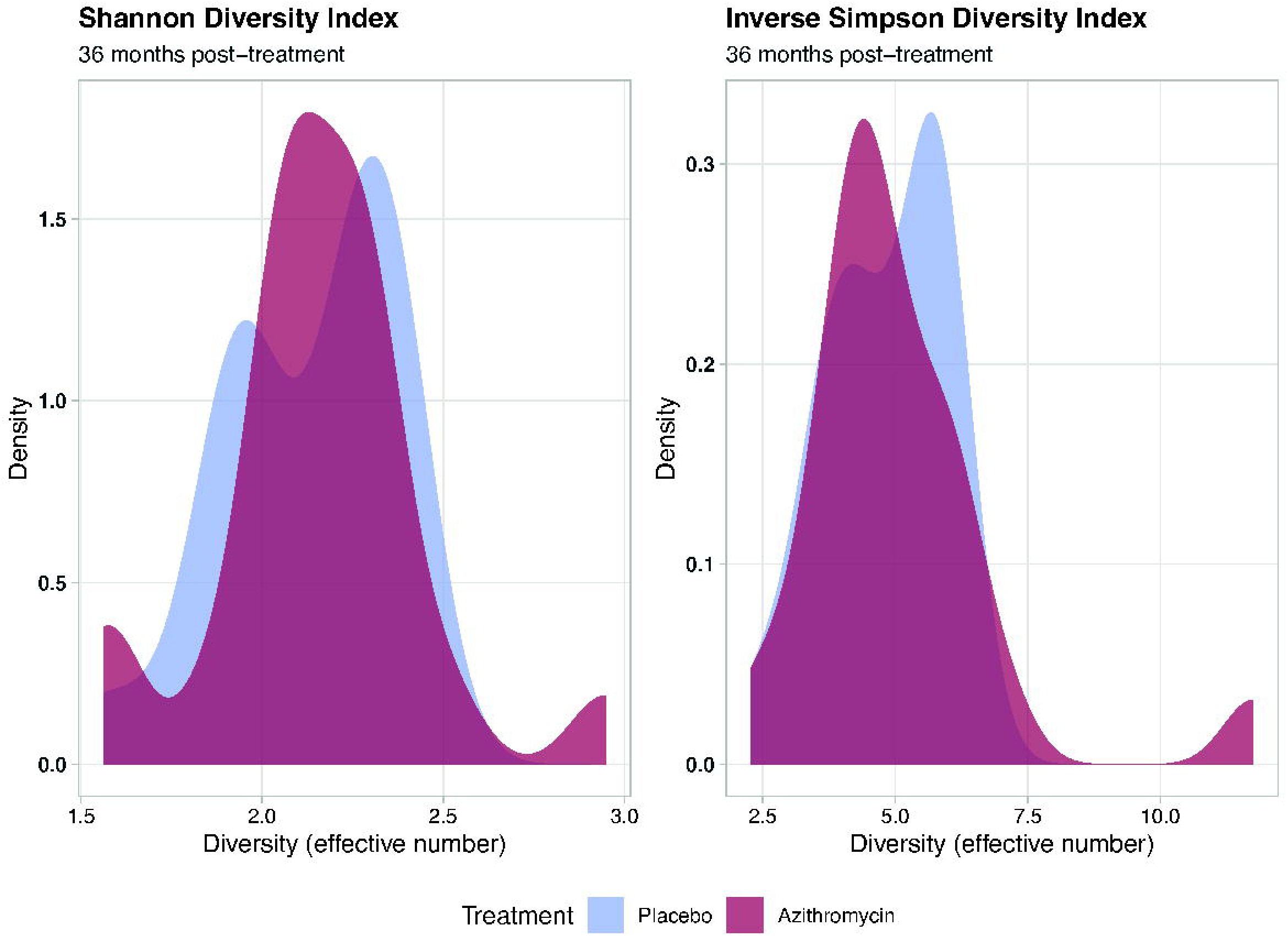
Shannon’s (A) and inverse Simpson’s (B) index for bacterial diversity at the genus level expressed in effective number in communities randomized to twice yearly azithromycin (purple) or placebo (blue) at 36 months.

## DISCUSSION

We did not find a significant increase in genetic resistance determinants to macrolide or any non-macrolide antibiotics in children 1-59 months old in communities that received 36 months of twice-yearly mass azithromycin distributions compared to placebo. While there were slight increases in macrolide resistance determinants in the azithromycin communities, the difference was not statistically significant and the magnitude of the effect was far smaller than has been observed in previous studies of mass azithromycin distribution in Niger.^9,17^ At 36 months in the MORDOR-Niger study, genetic macrolide resistance determinants were 7.4 times as high in azithromycin communities as compared to placebo.^9^ While reasons for a smaller increase in macrolide resistance in this setting compared to MORDOR-Niger are unclear, factors such as differences in background antibiotic use or SMC distribution may contribute to the differences observed in the present study in Burkina Faso compared to Niger. Previous work in Burkina Faso has demonstrated that antibiotic prescription is common in children under age 5 in Nouna, and that erythromycin is prescribed for some conditions.^18,19^ Increased background antibiotic use could mask differences between treatment groups, leading to only a modest increase in selection for antibiotic resistance due to MDA.

MORDOR-Niger found a statistically significant increase in selection for beta-lactam resistance in communities receiving azithromycin at 36 months, but no evidence that this was sustained at 60 months.^9^ Beta-lactams are one of the most commonly prescribed antibiotic classes for children in rural sub-Saharan Africa.^18–20^ Given the importance of this antibiotic class therapeutically, selection for beta-lactam resistance is an important outcome to monitor in mass azithromycin distribution programs. Here, we found no evidence of an increase in beta-lactam resistance after 36 months of twice-yearly distributions. Although continued monitoring as programs scale up is warranted, this finding is reassuring that in settings with similar antibiotic use patterns, distribution of azithromycin for prevention of child mortality may not select for beta-lactam resistance.

Previous work at the individual level has documented short-term changes in the composition of the microbiome following azithromycin administration, with no evidence of longer-term changes.^21–23^ Among infants in Pakistan, a single dose of azithromycin led to increased *Bifidobacterium infantis* colonization and reduced bacterial enteropathogens 56 days after birth.^24^ Intrapartum azithromycin has also been shown to affect the development of the microbiota of the gut of infants.^25^ At the community level, alpha diversity of the gut microbiome has generally been similar in communities receiving azithromycin compared to placebo, although evidence suggests that there may be differences in genera of bacteria, including reductions in potentially pathogenic bacteria such as *Campylobacter* spp.^26–29^ This study contributes to the evidence that azithromycin distribution does not alter the gut microbiome of children at the community level.

There are several limitations to this analysis. The evolving insecurity situation in Burkina Faso meant that some clusters that were included at baseline were no longer available at 36 months, and not all selected clusters at baseline could be sampled. While we replaced clusters at 36 months to increase the sample size, these replacement clusters did not have baseline measurements. Furthermore, since fewer clusters were sampled than originally planned, statistical power was lower than planned. However, point estimates for fold changes in antibiotic resistance determinants were close to the null, and were substantially smaller than those from previous studies. There was a chance imbalance in macrolide resistance determinants at baseline, with a higher read count of macrolides in the azithromycin compared to the placebo group. As previously noted, baseline measurements were not available for all clusters sampled at 36 months. However, if by chance antibiotic use was higher in the azithromycin clusters, we would expect this chance imbalance to lead to estimates that were further from the null at 36 months and thus do not explain the lack of increase in antibiotic resistance in azithromycin compared to placebo communities. Finally, the generalizability of these results is likely limited to areas that have similar background antibiotic use patterns.

Overall, these results suggest that twice yearly distribution of azithromycin to children aged 1-59 months did not lead to selection for antibiotic resistance in Burkina Faso over a 36 month period. Multiple trials have demonstrated the benefits of mass azithromycin distribution to children ages 1-59 months for the prevention of child mortality.^1–3^ While ongoing monitoring of antibiotic resistance in the context of programmatic scale-up will remain an important component of azithromycin implementation, these results are reassuring that azithromycin distribution for prevention of child mortality will not lead to substantial increases in antibiotic resistance in settings similar to the present trial.

## Data Availability

All data produced in the present study are available upon reasonable request to the authors

